# A Decade of Clinical Experience with DARPins in Oncology and Virology: A Systematic Review

**DOI:** 10.1101/2025.05.21.25328103

**Authors:** Michael T. Stumpp, Vladimir Kirkin, Michael P. Sanderson, Keith M. Dawson, Philippe Legenne

## Abstract

DARPins (Designed Ankyrin Repeat Proteins) are a non-antibody based protein scaffold which has undergone extensive clinical characterization. DARPins are small, stable proteins derived from naturally occurring ankyrin repeat proteins and are characterized by high binding affinity and specificity, and a flexible architecture enabling multispecific approaches beyond bi-specific molecules. A total of seven clinical DARPin drug candidates across therapeutic areas spanning ophthalmology (abicipar), virology (ensovibep), and oncology – including solid tumors (MP0250, MP0274, MP0310, MP0317) and hematologic neoplasms (MP0250, MP0533) – have been tested in patients, with the two most advanced reaching the registrational phase of development. Here, we systematically review the published clinical experience with DARPin therapeutics and imaging agents generated over the past decade. The DARPin drug candidates have evolved from relatively simple binders/neutralizers, with impact on specific disease hallmarks, to more sophisticated effector function-enabled molecules that modulate complex tumor-host interactions with therapeutic intent. The accumulated clinical safety, pharmacokinetics, and activity data described herein illustrate the versatility of DARPins to influence complex biological processes and thus help patients with medical conditions that are in some cases difficult to treat with other therapeutic modalities. Following recent breakthroughs in protein structure prediction and *de novo* protein design, DARPins are poised to deliver an array of unprecedented and exquisitely targeted novel drug candidates eagerly awaited by patients with high unmet medical needs.

## Introduction

Antibodies effectively link the adaptive and innate immune systems – a mechanism successfully exploited by modern therapeutic antibodies. Nevertheless, there is an ongoing quest to evolve beyond classical antibody-based therapeutics by endowing them with new functions. Bi-specific antibodies have been used in different therapeutic applications, most notably in oncology to stimulate an anti-tumor immune response in a targeted manner. As such, T cell engagers (TCEs) represent a widely utilized bi-specific antibody approach to direct T cell activation towards tumor cells with enriched expression of a single specific tumor-associated antigen (TAA) (Shouse, 2024). However, extension beyond bi-specific targeting, intrinsic to the bivalent immunoglobulin scaffold, has to date yielded limited success and many tumors lack a single “clean” TAA to target. Designed ankyrin repeat proteins, known as DARPins, represent a unique alternative to overcome the current limitations of therapeutic antibodies. This class of non-antibody protein scaffolds based on naturally occurring ankyrin repeat proteins has been studied extensively and developed as drug candidates over the past two decades (Plückthun, 2015; Stumpp, Dawson, and Binz, 2020). The small and stable nature of DARPin domains makes them amenable to assembly in linked peptide chains encoded by one gene and mimics familiar immunoglobulin functionalities. Like antibodies, DARPins can be selected for high affinity/potency and specificity towards diverse therapeutic targets. Distinct from antibodies, DARPins therapeutics can be designed with multiple specificities using up to six DARPin domains (this is the highest number of DARPin domains in one molecule tested to date). By including one or more DARPin domains targeting human serum albumin (HSA), the pharmacokinetic properties of these drugs can be tuned to enable the desired half-life, and thus receptor occupancy, for a given therapeutic modality without using the Fc-region of antibodies. The manufacturing of DARPin therapeutics relies on *Escherichia coli* as the expression host, followed by chromatographic purification – a simple and scalable process, which expedites clinical development.

Previous review articles have comprehensively described the structural and functional features of DARPin-based drugs (Stumpp et al., 2020), as well as the collective clinical data from the first DARPin therapeutic abicipar pegol (MP0112; AGN-150998); a half-life extended VEGF-A inhibitor for intraocular application in wet age-related macular degeneration (AMD) (Desideri, Traverso, and Nicolò, 2020); **Box**). In this article, we review the properties of the six systemically administered DARPin drug candidates that have been tested over the last decade in oncology and virology clinical settings, as well as recent clinical experience with radiolabeled DARPins being developed for imaging purposes.

#### Box

Abicipar pegol, the first DARPin studied clinically, is a one-domain anti-VEGF-A DARPin carrying a 20 kDa polyethylene glycol (PEG) moiety to extend the local residence time in the vitreous body of patients with ocular diseases, previously reviewed in (Desideri et al., 2020) and (Stumpp et al., 2020). Clinical development mainly focused on patients with wet AMD. In these patients, abicipar dosed every 8–12 week demonstrated non-inferiority in two phase 3 studies in terms of stable vision and central retinal thickness when compared to monthly treatment with ranibizumab (an approved anti-VEGF-A antibody developed by Genentech, Inc.). However, increased rates of intraocular inflammation were reported in about 15% of patients treated with abicipar (Khurana et al., 2021; Kunimoto et al., 2020). Subsequent manufacturing changes reduced this inflammation to the upper single-digit percent range (Callanan et al., 2023), and recent evidence suggests that part of the inflammation also observed with other ocular protein drugs was likely caused by changes induced by silicone oil contained in syringes (Anderson et al., 2021; Melo et al., 2021). Therefore, using silicone oil-free syringes represents a potential avenue for future exploration for abicipar.

## Methods

### Information Sources & Search Strategy

A systematic literature review was conducted based on PubMed searches, and relevant publications from 01 Jan 2014 to 15 Apr 2025 were included. The search strategy is listed in **Table 1**. In addition, pragmatic searches were performed using Google and websites of relevant scientific societies for related congress abstracts (2021–2024; **Table 1**). Reference lists of retained publications were searched for additional relevant sources (“snowballing”).

**Table 1.** Systematic search strategy.

| LITERATURE TYPE | SEARCH STRATEGY | NO. OF IDENTIFIED SOURCES |
| --- | --- | --- |
| **PUBMED**<br>**01 JAN 2014 –**<br>**15 APR 2025** | #1. (“DARPin” OR “designed ankyrin repeat protein”) filtered for article type “Clinical trial” | 8 |
|  | #2. (“DARPin” OR “designed ankyrin repeat protein”) AND (clinical study OR clinical trial) | 23 |
|  | #3. (“DARPin” OR “designed ankyrin repeat protein”) AND (“phase 1” OR “phase 2” OR “phase 3”) | 27 |
| **CONGRESS ABSTRACTS**<br>**01 JAN 2021–15**<br>**APR 2025\*** | Congress abstract books of AACR, ASCO, ASH, EHA and SITC were searched using the search terms “DARPin” and “Designed Ankyrin Repeat Protein” | 36 |
| **TOTAL** |  | **94** |

Abbreviations: AACR, American Association for Cancer Research; ASCO, American Society of Clinical Oncology; ASH, American Society of Hematology; DARPin, Designed Ankyrin Repeat Protein; EHA, European Hematology Association; SITC, Society for Immunotherapy of Cancer
\*Includes 2021-2025 abstracts for AACR, and 2021-2024 abstracts for ASCO, ASH and EHA due to their respective congress timings.

### Eligibility Criteria

Clinical studies of all phases reporting results of DARPins in oncology or virology were included. Exclusion criteria were clinical studies in other indications, review articles, and articles not written in English.

### Study Selection & Data Collection

Studies were selected and data collected by three authors (MPS, MTS, and VK) and quality checked by two authors (KMD and PL). The collected data contained information on the study design (phase), population (number of patients, indication) as well as primary and secondary study outcomes, such as safety, efficacy, and PK/PD assessments. The extracted data are reported descriptively, no meta-analysis or other data synthesis was performed.

## Results

### Study Selection & Characteristics

A total of 94 published sources were identified (**Table 1**). After removal of duplications, 72 sources were screened for eligibility, including 36 full articles and 36 congress abstracts. Of the 15 eligible and reviewed sources, 11 were retained (eight full articles and three congress abstracts). With snowballing, another two published articles relevant for this systematic review were identified and included (**Figure 1**). Thus 13 published sources identified for this systematic review reported results of five DARPins evaluated as therapeutics (four in oncology and one in virology) and two DARPins assessed for imaging in oncology (**Table 2**). In addition, data on file available to the authors for another DARPin therapeutic with an oncology indication was included from a completed but not yet published clinical trial (**Table 2**).

**Figure 1.**
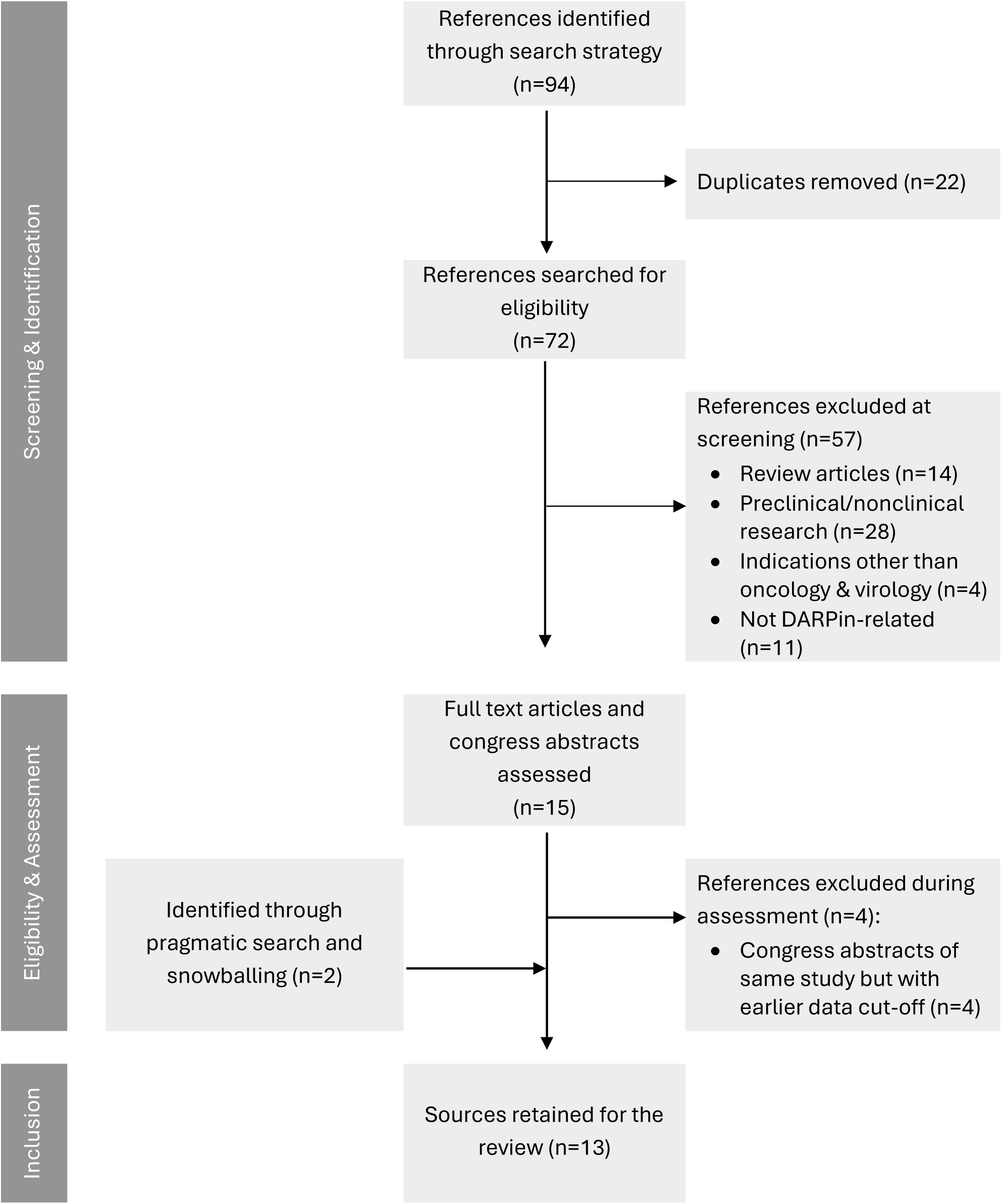
Flow chart for the identification and selection process of published sources included for this systematic review.

**Table 2.** Overview of clinical studies with DARPins in oncology and virology patients*.

| DARPIN | REFERENCE IDENTIFIED | PHASE | INDICATION | PATIENTS TREATED | DOSE RANGE TESTED | CLINICAL TRIAL IDENTIFIER |
| --- | --- | --- | --- | --- | --- | --- |
| **MP0250** | (Baird et al., 2020) | 1 | Advanced solid tumors | 45 | 0.5 – 12 mg/kg | NCT02194426 |
|  | (Knop et al., 2024) | 1b/2 | Multiple Myeloma | 33 | 8 – 12 mg/kg | NCT03136653 |
|  | Data on file, Molecular Partners AG | 1b/2 | NSCLC | 8 | 8 mg/kg | NCT03418532 |
| **MP0274** | (Fischer et al., 2022) | 1 | HER2-positive solid tumors | 22 | 0.5 – 12 mg/kg | NCT03084926 |
| **MP0310** | Data on file, Molecular Partners AG | 1 | Advanced solid tumors | 37 | 0.015 – 12 mg/kg | NCT04049903 |
| **MP0317** | (Krieg et al., 2024; Steeghs et al., 2024) | 1 | Advanced solid tumors | 46 | 0.03 – 10 mg/kg | NCT05098405 |
| **MP0420 / ENSOVIBEP** | (Prins et al., 2022) | 2a | Non-hospitalized COVID-19 | 12 | 225 mg & 600 mg | NCT04834856 |
|  | (ACTIV-3/TICO_Study_Group, Barkauskas, et al., 2022) | 2 | Hospitalized COVID-19 | 247 | 600 mg | NCT05780463 |
|  | (Kingsley et al., 2024) | 2 | Non-hospitalized COVID-19 | 301 | 75 mg, 225 mg & 600 mg | NCT04828161 |
| **MP0533** | (Jongen-Lavrencic et al., 2024) | 1/2a | AML, MDS | 37<sup>†</sup> | Undisclosed, 7 dose levels | NCT05673057 |
| <sup>99m</sup>**TC-HE<sub>3</sub>-G3** | (Bragina et al., 2022) | 0 | HER2 imaging | 28 | 1-3 mg | NCT04277338 |
| <sup>99m</sup>**TC-HE<sub>3</sub>-G3** | (Bragina et al., 2023) | 0 | HER2 imaging | 11 | 3 mg | NCT05376644 |
| <sup>99m</sup>**TC-HE<sub>3</sub>-EC1** | (Zelchan et al., 2024) | 0 | EpCAM imaging | 12 | 3 mg | NCT05620472 |

\*One additional study in healthy volunteers was identified in the systematic literature review (Stojcheva et al., 2023): a phase 1 study in 17 individuals treated with ensovibep 3, 9 & 20 mg/kg (NCT04870164). The results of this study are also discussed in detail below.
<sup>†</sup>Study still ongoing, dose ranges 1–7 recruited 37 patients, interim data shown at ASH 2024.

The following DARPin molecule structure images were published previously (all under <u>CC BY-NC-ND 4.0</u>): MP0250 (Knop et al., 2024), MP0310 (Saltarella et al., 2024) and MP0533 (Bianchi et al., 2024).

### MP0250 (VEGF x HGF) – a dual inhibitor for patients with solid tumors

The first DARPin drug candidate developed for systemic administration was MP0250, a multi-specific DARPin designed for dual-inhibition of the growth factors VEGF-A and HGF (Binz et al., 2017; Fiedler, Ekawardhani, et al., 2017). The rationale behind the design of MP0250 was that many tumors rely on VEGF-A and HGF, derived either from autocrine expression or from other cells within the tumor microenvironment (TME) niche, to support their growth, survival, invasion into healthy tissue, and drug resistance (Saltarella et al., 2023). To ensure durable suppression of VEGF-A and HGF signaling, MP0250 was designed to have a long half-life via the inclusion of two HSA binding DARPin domains (Steiner et al., 2017). As a result, MP0250 is composed of four functional protein domains (2x anti-HSA, 1x anti-VEGF-A and 1x anti-HGF; Figure 2).

**Figure 2.**
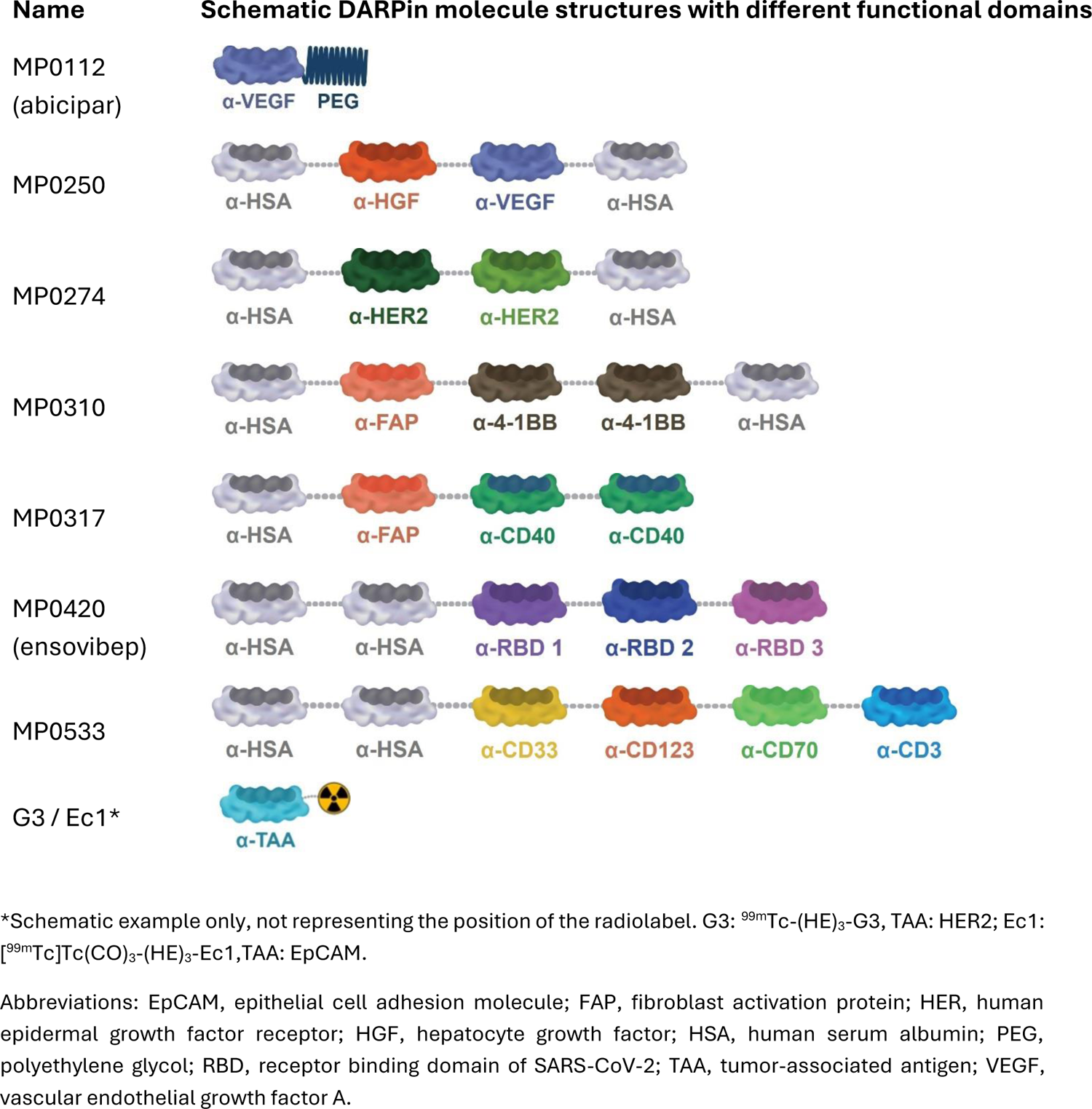
Overview of the DARPin drug candidate structures.

A dose-escalation phase 1 study in patients with advanced solid tumors was initiated in 2014 (Baird et al., 2020), and recruited a total of 45 patients across 7 planned dose cohorts (5 dose levels and 2 additional dose cohorts at a different dosing interval and duration of infusion, NCT02194426, **Table 2**). Based on data from preclinical models (Fiedler, Ekawardhani, et al., 2017), the clinical starting dose was determined to be 0.5 mg/kg, applied intravenously (i.v.) every other week. Dose-limiting adverse events of MP0250 were reported to be hypertension and proteinuria, consistent with inhibition of the VEGF-A pathway (Baird et al., 2020). Hypoalbuminemia was reported in about a quarter of patients, which has also been described for other agents targeting the HGF axis (Cortot et al., 2022). The median number of infusions was four, with some patients receiving treatment for more than one year (and in one patient for 60 weeks, corresponding to 31 infusions). One unconfirmed partial response (PR) was reported as the best overall response. About one third of the patients had a reduction of tumor lesion size from baseline. In keeping with the half-life extension properties of the MP0250 molecule design, the maximum terminal half-life observed ranged from 15 to 16 days (depending on the method of half-life calculation across all dose cohorts), allowing for convenient dosing every 2–4 weeks. Consistent with the desired mechanism of action, MP0250 led to a reduction of free VEGF-A in plasma to undetectable levels. A dose-dependent elevation of total plasma HGF levels was observed, reflecting an increased abundance of MP0250-HGF complexes. Anti-drug antibodies (ADAs) were observed in about half of the patients and had no discernible effects on MP0250 exposure, biological activity, or safety outcomes. The determined recommended phase 2 doses were 8 mg/kg every 2 weeks (Q2W) or 12 mg/kg every 3 weeks (Q3W).

This phase 1 study represented a significant milestone for the application of DARPin therapeutics in oncology. The acceptable safety profile, extended pharmacokinetics, and desired biological activity of MP0250, coupled with flexible dosing options and negligible impact of ADAs, provided a strong impetus for the initiation of two phase 2 trials of MP0250 in combination with approved standard of care treatments.

The first phase 1b/2 study, initiated in 2017, combined MP0250 with the proteasome inhibitor bortezomib and dexamethasone in patients with relapsed or refractory (r/r) multiple myeloma (MM) (NCT03136653; (Knop et al., 2024)). This clinical setting was selected based on the established roles of VEGF-A and HGF in promoting tumor growth, survival, angiogenesis, and drug resistance in MM (Ferrucci et al., 2014; Zhang, Su, Volpert, and Woude, 2003), as well as preclinical data demonstrating the utility of MP0250 in blocking angiogenesis in translational models of human MM (Rao et al., 2018). In this open-label single-arm study, a total of 33 patients with r/r MM were treated with MP0250 at dose levels up to 12 mg/kg Q3W. In part 1 (11 patients), two dose-limiting toxicities (DLTs) were reported and the maximum tolerated dose (MTD) was therefore set at 8 mg/kg Q3W for part 2 (22 patients). Objective responses were reported in 32% of patients (9/28 patients evaluable for response) (Knop et al., 2024). Responders had a median duration of response (mDoR) of 8 months and a median progression-free survival (mPFS) of 4.2 months. The two patients with the longest treatment duration were shown to have sustained full MP0250 exposure for the entire treatment duration of 20 and 30 months, respectively. The most frequently reported treatment-related adverse events (TRAEs) were hypertension (58% of patients), thrombocytopenia (32%), proteinuria (29%), and peripheral oedema (19%). Consistent with data from the phase 1 study described above (Baird et al., 2020), the steady-state terminal half-life of MP0250 was around 15 days. ADAs were detected in 13% of patients, with no discernible impact on MP0250 exposure, biological activity, safety, or efficacy.

In conclusion, MP0250 in combination with bortezomib and dexamethasone showed promising clinical activity and acceptable safety in r/r MM patients. These findings suggest that dual-targeting of HGF and VEGF-A signaling, in combination with proteasome inhibition and dexamethasone, represents an attractive therapeutic strategy in MM, and provide an impetus for deeper analysis into the molecular features of patients who will derive the greatest benefit from this approach. Despite this, as a consequence of the advent of novel and highly active drugs in the MM armamentarium, namely agents targeting CD38, SLAM family member 7 (SLAM7), or B cell maturation antigen (BCMA) (Lin et al., 2024), the clinical development of MP0250 in MM was not pursued further.

The second phase 1b/2 study, initiated in 2018, combined MP0250 with the third-generation epidermal growth factor receptor (EGFR) inhibitor osimertinib in patients with EGFR-mutated non-small cell lung cancer (NSCLC), who had documented progression on osimertinib (NCT03418532). The rationale for this study was the emerging evidence that the HGF signaling pathway played a significant role in acquired resistance to osimertinib in NSCLC patients in the AURA3 clinical trial (Papadimitrakopoulou et al., 2018). This open-label single-arm study enrolled a total of eight patients but was ultimately terminated before the phase 2 part commenced (study sponsor’s decision). The decision to focus development of MP0250 in MM was made in the context of the entire MP0250 development program and partly informed by the observation of an increased incidence of adverse events related to VEGF-A inhibition in the NSCLC patients treated with 8 mg/kg Q3W compared to other MP0250 studies (data on file, Molecular Partners AG). Given the recent clinical success of targeting the HGF receptor (c-MET) to overcome acquired resistance to EGFR inhibitors (Wu et al., 2024), further examination of the utility of MP0250 at a lower dose could be warranted to address the high therapeutic need of EGFR-mutated NSCLC patients.

### MP0274 (HER2 x HER2) – a bi-paratopic, pro-apoptotic inhibitor for patients with HER2-positive tumors

The second systemically administered multi-specific DARPin drug candidate was MP0274, which was designed to include two DARPin domains for bi-paratopic targeting of distinct epitopes of the human epidermal growth factor receptor 2 (HER2/ERBB2), as well as two HSA-binding DARPin domains for half-life extension (Figure 2; (Fiedler, Metz, et al., 2017)). Genomic amplification and overexpression of HER2 occurs in a subset of patients with breast cancer, and has led to the successful development of diverse HER2-targeting agents (Stebbing, Copson, and O’Reilly, 2000). Beyond the established oncogenic role of HER2, the rationale for pursuing clinical development of MP0274 was supported by preclinical data demonstrating a differentiated mechanism of action of bi-paratopic HER2 binding, compared to the approved mono-paratopic HER2 binders trastuzumab and pertuzumab (Dolado et al., 2013; Mitra et al., 2017). Only bi-paratopic HER2-binding DARPins, like MP0274, were able to induce apoptosis in HER2-expressing cancer cells *in vitro*. Additionally, MP0274 delivered superior *in vivo* efficacy in HER2-expressing patient-derived xenograft (PDX) models compared to trastuzumab or pertuzumab monotherapy, and non-inferiority compared to the combination of trastuzumab with pertuzumab (Fiedler, Metz, et al., 2017). Direct tumor cell killing independent of antibody-dependent cellular cytotoxicity (ADCC) or antibody-dependent cellular phagocytosis (ADCP) was hypothesized to offer a distinct therapeutic mechanism compared to antibody-based HER2-targeting drugs, particularly in patients displaying suppressed immune function.

The MP0274 phase 1 single arm open label study (NCT03084926) was initiated in 2017 (Fischer et al., 2022). A total of 22 patients with advanced/metastatic solid tumors with histologically confirmed tumor expression of HER2 were enrolled in the study, including patients who had previously received HER2-targeting agents. Patients received MP0274 at six dose levels between 0.5 to 12 mg/kg i.v. Q3W (**Table 2**). MP0274 was generally well tolerated. The most frequent TRAEs were infusion-related reactions (IRRs) observed across all dose levels (in 7 out of 22 treated patients), mostly grade 2 and including one grade 3 event. PK analyses indicated the anticipated target-mediated drug disposition (TMDD) for HER2-targeting biologics. Thus, the terminal half-life was estimated at around 15 days, based on PK data from the highest tested doses. In total, about half of the patients (12/22) were reported to have ADAs in at least one sample. Stable disease was reported in three patients, at 0.5, 4, and 8 mg/kg, lasting 8.8, 4.7, and 4.4 months, respectively. The best response was a PR lasting for 17 months in a patient with breast cancer treated with MP0274 at 12 mg/kg. This patient displayed sustained MP0274 exposure, and no ADAs were detected. Of note, this patient had received prior treatment with the combination of trastuzumab, pertuzumab, and docetaxel, as well as later treatment with trastuzumab emtansine.

In conclusion, this phase 1 study with MP0274 was successfully completed and provided an amenable path for follow-up clinical studies in HER2-overexpressing cancer settings. Nevertheless, akin to the experience with MP0250, the emergence of novel and highly active HER2-targeting modalities, principally the antibody drug conjugate (ADC) fam-trastuzumab deruxtecan-nxki (T-Dxd/ENHERTU®) (P. Narayan et al., 2020), ultimately influenced the decision not to further pursue clinical development of MP0274.

### MP0310 (FAP x 4-1BB) – a FAP-localized, 4-1BB agonistic co-stimulatory immune cell engager for patients with solid tumors

Based on the encouraging initial clinical data for the first two DARPin drug candidates in oncology, MP0250 and MP0274, and the advent of diverse immune-oncology therapies at the time, the amenability of DARPins for multi-specificity was directed towards the design of more sophisticated immune modulators. Specifically, MP0310 constitutes the first bi-specific agonistic, immune-modulatory DARPin designed for tumor-localized co-stimulation of immune cells (Claus, Ferrara-Koller, and Klein, 2023). MP0310 contains two adjacent agonistic DARPin domains (Figure 2) binding the TNF receptor superfamily member 4-1BB (also called CD137); a co-stimulatory factor endogenously expressed on T cells and NK cells (Chester, Sanmamed, Wang, and Melero, 2018).

MP0310 also includes a single DARPin domain binding the type II transmembrane serine protease fibroblast activation protein (FAP), which is highly expressed by tumor stromal cells across diverse cancer indications (Dziadek et al., 2024). This design aimed to achieve localized activation of the 4-1BB pathway within the TME. Preclinical studies with MP0310 confirmed a potent co-stimulatory effect on T cells only in the presence of FAP; an effect which significantly inhibited the growth of tumor xenografts in humanized mouse models (Link et al., 2018; Saltarella et al., 2024). In contrast to systemic 4-1BB agonistic antibodies, MP0310 did not induce hepatotoxicity or systemic T cell activation manifesting in graft versus host disease (GvHD). Importantly, in healthy cynomolgus monkeys, MP0310 did not provoke memory T cell proliferation despite being fully cross-reactive with cynomolgus 4-1BB and FAP (data on file, Molecular Partners AG). This result indicated that the localized activation of 4-1BB by MP0310 strictly depends on FAP-mediated clustering, which is absent in healthy animals.

Phase 1 clinical development of MP0310 in advanced solid tumor patients started in 2019 (NCT04049903). The phase 1 starting dose was chosen in accordance with the minimum anticipated biological effect level (MABEL) approach (Link et al., 2020), and a total of 37 patients were treated in seven increasing dose cohorts of MP0310 administered i.v. Q3W (**Table 2**). Additionally, two cohorts received MP0310 weekly (Q1W, at different dose levels), and one cohort was pretreated with the anti-CD20 antibody rituximab prior to Q3W dosing in an effort to mitigate the risk of ADAs (see below). The most frequent TRAEs were IRRs, observed in 12/22 patients in the Q3W cohort, predominantly at higher doses, and in 3/9 patients in the Q1W cohorts (manuscript in preparation). IRRs typically recurred upon re-exposure. MP0310 was not associated with signs of autoimmune reactions. Further common treatment-emergent adverse events (TEAEs) were fatigue and nausea. The best overall response to MP0310 monotherapy was one unconfirmed PR in a patient with melanoma, whilst stable disease was observed in 16 (43%) patients. The terminal half-life of MP0310 was between 1–2 weeks depending on the dose level, consistent with the inclusion of two HSA-binding DARPin domains in the molecule (Figure 2). Similar to MP0274, TMDD was observed at low doses of MP0310, as anticipated. Most patients developed ADAs which frequently coincided with a reduction of MP0310 exposure after the first treatment cycle. Of interest, clinical studies with 4-1BB agonistic antibodies including urelumab (Segal et al., 2018; Timmerman et al., 2020) and cinrebafusp alfa (Piha-Paul et al., 2024) have also reported a relatively high incidence of ADAs. The emergence of ADAs against 4-1BB agonists may in part reflect the uniquely broad costimulatory function of 4-1BB on multiple lymphoid lineages (B, NK, and T cells) and antigen presenting cells (APCs) such as macrophages and dendritic cells (DCs), which upon activation, may potentiate the humoral immune response for ADA generation (Yanchen Zhou et al., 2022). Importantly, the observed ADAs did not change the safety profile of MP0310, and an MTD was not established within the dose range tested.

Further development of MP0310 was not subsequently pursued despite the acceptable safety profile and early signs of clinical activity in this phase 1 study. Nevertheless, the data provided valuable insights into the potential of tumor-localized 4-1BB agonists. The clinical development of multiple systemic 4-1BB agonist antibodies has been hampered by pronounced safety concerns including hepatotoxicity (Singh, Kim, Lee, Eom, and Choi, 2024). Akin to the experience with MP0310, a phase 1 dose-escalation study of RO7122290, a fusion protein of a split trimeric 4-1BB ligand and a fragment antigen-binding (Fab) moiety targeting FAP, reported an acceptable safety profile and did not identify an MTD (Melero et al., 2024). These findings suggest that utilization of FAP, or other tumor antigens, for localized activation of 4-1BB is a feasible path towards mitigating safety risks associated with systemic 4-1BB pathway activation. In addition, the bispecific antibody acasunlimab, displaying conditional 4-1BB activation upon PD-L1 binding on tumor or immune cells, in combination with the anti-PD-1 antibody pembrolizumab, was recently reported to deliver promising efficacy in metastatic NSCLC patients, including those previously treated with pembrolizumab (Aerts et al., 2024). Together, these findings indicate that utilization of tumor-associated antigens (TAAs), such as FAP, for localized 4-1BB activation represent an attractive therapeutic approach to enhance antitumor immunity and deliver a therapeutic benefit to patients.

### MP0317 (FAP x CD40) – a FAP-localized, CD40 agonistic co-stimulatory immune cell engager for patients with solid tumors

The second agonistic, immuno-modulatory DARPin drug candidate to enter clinical development was MP0317. Like MP0310, MP0317 utilizes the tumor-localizing principle based on FAP-binding. However, unlike MP0310, the target immune cells are APCs expressing CD40. MP0317 relies on FAP-binding to induce the clustering of CD40 on APCs via two CD40-binding DARPin domains (Figure 2) and leads to specific activation of B cells, DCs, and macrophages *in vitro* as evidenced by the upregulation of cell surface activation markers and release of cytokines (Rigamonti et al., 2022). *In vivo*, a surrogate DARPin with potent binding of murine FAP and CD40 induced complete regression in subcutaneous FAP-expressing MC38 colorectal tumors xenografted to immune competent mice, as well as protection against rechallenge with parental or FAP-expressing MC38 cells. In contrast to systemic CD40 agonists, the surrogate of MP0317 did not cause an elevation of liver enzymes or systemic cytokine release in mice, supporting the therapeutic concept of tumor localized CD40 activation.

A phase 1 dose escalation study in patients with advanced solid tumors was initiated in 2021 and recruited a total of 46 patients (NCT05098405) (Krieg et al., 2024; Steeghs et al., 2024). Nine dose cohorts of MP0317 (0.03 – 10 mg/kg, based on MABEL considerations) were tested, which were administered i.v. either Q1W or Q3W. Eligible patients with advanced solid tumors were selected from indications previously published to display high tumor FAP expression, with colorectal cancer (CRC) representing the most prevalent tumor type (12/46 or 26% of patients) in the study.

MP0317 demonstrated a favorable safety profile; the most frequent TRAEs were fatigue and IRRs (Steeghs et al., 2024). A single patient receiving the highest MP0317 dose level (10 mg/kg, Q3W) experienced a DLT of asymptomatic transient grade 3 alanine and aspartate aminotransferases elevation. Based on the totality of safety data, an MTD was not reached. The best response to MP0317 monotherapy was an unconfirmed PR in a patient with gastrointestinal stromal tumor (GIST). A further 14 (30%) patients achieved stable disease(Steeghs et al., 2024). The PK profile of MP0317 was consistent with the half-life extended properties enabled by the inclusion of a single HSA binding DARPin domain in the molecule (Figure 2). At higher MP0317 dose levels, exposure and pharmacodynamic activity (see below) were sustained over multiple cycles. This reflected the ability of these dose levels to overcome the impact of ADAs, observed in a majority of patients, and TMDD.

To confirm the desired mechanism of action of MP0317, an extensive biomarker analysis package was performed using both paired tumor biopsies and peripheral samples (Krieg et al., 2024). Tumor localization of MP0317 was confirmed, which was associated with pronounced TME remodeling consistent with activation of the CD40 pathway. At higher MP0317 dose levels, the Q1W and Q3W schedules both led to increased DC abundance and maturation in the TME. Additional TME remodeling by MP0317 was exemplified by increased abundance of M1 macrophages, plasma cells, and T follicular helper cells, as well as activation of IFNγ downstream signaling. Peripheral PD effects were monitored longitudinally and included an increase in the CXCL10 chemoattractant, a transient reduction of peripheral B cell abundance, and activation of immune cells. Importantly, the CD40 pathway was activated by MP0317 in a broad-spectrum of cancer types and various tumor locations.

The relatively modest clinical efficacy of MP0317 monotherapy mirrors the outcomes reported with other CD40 agonists (Jian et al., 2024; Yang Zhou, Richmond, and Yan, 2023). These findings suggest that the greatest value of CD40 agonist agents will be realized in combination regimens, particularly with agents that complement the TME remodeling induced by CD40 pathway activation. The favorable safety profile of MP0317, in contrast to systemic CD40 agonist antibodies, as well as flexibility to utilize different MP0317 dosing schedules, offer a unique opportunity for diverse drug combination approaches. In keeping with this, phase 2 drug combination studies in selected indications are currently being assessed.

### MP0420 (ensovibep; SARS-CoV-2 spike protein) – antiviral tri-specific with broad coverage of COVID-19 variants

The global emergence of COVID-19 in 2020 was an impetus for the development of diverse vaccines and therapeutic agents (e.g., neutralizing antibodies, small molecule viral protease inhibitors) against the SARS-CoV-2 virus. The receptor binding domain (RBD) of the SARS-CoV-2 spike protein represents the main target for therapeutic antibodies and vaccines and is prone to rapid antigen drift to create new virus variants that escape targeting (Yewdell, 2021). To address this limitation, the multi-specific DARPin molecule ensovibep (MP0420) was designed to include three adjacent anti-SARS-CoV-2 DARPin domains with slightly different paratopes, in order to ensure high-affinity binding to as many virus variants as possible and potentially counter antigen drift (Figure 2; (Walser, Mayor, and Rothenberger, 2022)). This was rapidly achieved by incorporating DARPin domains with slightly different sequences from the primary selection output, each of them with very high affinity.

Clinical development of ensovibep started in healthy volunteers in November 2020 (NCT04870164; (Stojcheva et al., 2023)), after only eight months of preclinical research and development to generate and characterize ensovibep. Pharmacodynamic insights were gained from a PK/PD model (Claas et al., 2024), and clinical doses as low as 75 mg were postulated to be effective in ambulatory adult patients with symptomatic COVID-19. In view of the acute clinical course of COVID-19 infection, and the expectation that ensovibep would display a long half-life in human, all clinical studies explored single ensovibep infusions. This provided valuable insights into the ensovibep PK profile, which confirmed the expected first-order pharmacokinetics over the time course of 13 weeks/91 days. The collective ensovibep clinical program comprised four clinical studies with a total of 560 patients, thus constituting the largest systemic DARPin dataset to date (**Table 2**).

Initial clinical data for ensovibep were collected in a single ascending dose study in a total of 17 healthy volunteers across three dose levels (NCT04870164; (Stojcheva et al., 2023)). Ensovibep was safe and well tolerated, and one adverse event of special interest (AESI) of moderate severity, hypersensitivity vasculitis, was reported at the 20 mg/kg dose (corresponding to 1200 mg in this patient). A terminal half-life of about 15 days was determined at the highest dose tested, and ensovibep clearance was mono-exponential. ADAs were observed in 15/17 healthy volunteers with limited initial impact on exposure, and with some accelerated ensovibep clearance being observed in a minority of healthy volunteers. *Ex vivo* analyses using pre- and post-treatment serum samples from the healthy volunteers on this study confirmed the potent and dose-dependent neutralization of SARS-CoV-2 virus by ensovibep.

The therapeutic activity of ensovibep in COVID-19 patients was first studied in a total of 12 patients at two different dose levels of 225 mg (n=6) and 600 mg (n=6), administered once via i.v. infusion (NCT04834856). This small, single-center phase 2a study was conducted between April and June 2021, at a time when the SARS-CoV-2 alpha strain was prevalent in the Netherlands (Vos et al., 2024). Similar to the experience in healthy volunteers, ensovibep was considered generally safe and well tolerated at both doses tested (Prins et al., 2022). The most notable TEAEs considered related to ensovibep treatment were diarrhea (2 patients) and elevated liver enzymes (3 patients). While a putative role of ensovibep could not be ruled out, these AEs could also have been related to the COVID-19 disease pathology. All patients remained on study for the entire study duration of 91 days. The terminal half-life of ensovibep was again reported in the range of two weeks, with 1/6 patients in each cohort showing accelerated clearance at about 3 weeks post infusion. ADAs were reported in 10/12 patients with no impact on ensovibep exposure during the acute phase of the infection (2 weeks). Viral titers in nasopharyngeal swabs declined rapidly, in line with the effects reported with monoclonal antibodies targeting SARS-CoV-2 (Weinreich et al., 2021). No differences were observed between the 225 mg and 600 mg doses, indicating that both doses of ensovibep achieved a maximal decrease of viral load.

As part of the global efforts to study and treat COVID-19, ensovibep was also included in the Accelerating COVID-19 Therapeutic Interventions and Vaccines (ACTIV-3)/Therapeutics for Inpatients with COVID-19 (TICO) international consortium protocol with the aim of treating hospitalized patients with COVID-19 (NCT04501978). A total of 247 patients were included in the ensovibep arm of this double-blind, randomized, placebo-controlled master protocol across 62 sites in 10 countries (ACTIV-3/TICO_Study_Group, Barkauskas, et al., 2022). An independent data safety monitoring board (DSMB) continuously reviewed the data and concluded that ensovibep combined with standard of care did not improve clinical outcomes for hospitalized COVID-19 patients. The ensovibep part of the protocol for recruitment was thus closed at an interim point for early futility. No new safety concerns were identified in this study. Of note, multiple other SARS-CoV-2 neutralizing biologics tested in the ACTIV-3/TICO study closed recruitment at an interim point for early futility (ACTIV-3/TICO_Bamlanivimab_Study_Group* et al., 2022; ACTIV-3/TICO_LY-CoV555_Study_Group et al., 2020; ACTIV-3/TICO_Study_Group, Self, et al., 2022). The neutralizing antibody combination tixagevimab-cilgavimab was the only agent reported to have proceeded to full enrolment. However, this treatment failed to improve the primary endpoint of sustained patient recovery (Holland et al., 2022). These findings suggest that agents neutralizing the SARS-CoV-2 virus may have limited application in hospitalized patients, who have relatively advanced disease and inflammation-related pathologies.

Finally, a large global phase 2 study with three dose levels of ensovibep (75 mg, n=103; 225 mg, n=102; 600 mg, n=100) and one control (placebo) cohort (n=102) was conducted in non-hospitalized COVID-19 patients (EMPATHY, NCT04828161; (Kingsley et al., 2024)). The study recruited from May to October 2021 at 48 sites in five countries across four continents, thus mostly including patients infected by the delta variant of SARS-CoV-2. The unique approach of this study was to test whether ensovibep doses as low as 75 mg, administered once by infusion (1 h), would lead to statistically significant and clinically relevant treatment benefits versus placebo. The primary endpoints were related to SARS-CoV-2 viral load (part A) and the rate of COVID-19-related hospitalizations, emergency room (ER) visits, or deaths from any cause (part B). The study reached the primary endpoint of time-weighted change from baseline in log_10_ SARS-CoV-2 viral load through day 8 at all dose levels tested. Additionally, ensovibep-treated patients showed fewer COVID-19-related hospitalizations, ER visits, and all-cause mortality compared to placebo, resulting in a relative risk reduction of 78%. No unexpected safety findings were reported, and PK analyses showed the expected half-life of close to two weeks. ADAs above threshold in a validated assay were reported in 55–75% of all patients but did not impact the safety or efficacy profile of ensovibep.

Despite the positive phase 2 data and differentiated therapeutic profile of ensovibep, the change in overall disease impact of the COVID-19 pandemic, coupled with ensovibep’s lower potency against new COVID-19 variants, led to a deprioritization of further clinical development. The EUA (emergency use authorization by Food and Drug Administration, FDA) for ensovibep was withdrawn effective January 2023 and clinical development was stopped. Nevertheless, a significant learning from the ensovibep program is that a highly specific and potent multi-specific antiviral DARPin could be developed from concept to first-in-human application in less than eight months; a feat enabled by the favorable developability profile of DARPins, and the special circumstances surrounding the global COVID-19 pandemic. Notably, a preclinical study showed that ensovibep remained fully potent across the early and late SARS-CoV-2 variants (e.g., wild-type, alpha, beta) and early omicron variants (Rothenberger et al., 2022). These data highlight the utility of DARPins for multi-specific targeting of viral variants and the potential mitigation of viral antigen drift.

### MP0533 (CD33 x CD123 x CD70 x CD3) – avidity-gated tetra-specific T cell engager for patients with AML

Acute myeloid leukemia (AML) is a highly heterogenous hematological disorder characterized by defective differentiation of the myeloid cell lineage leading to production of leukemic stem cells (LSCs) and proliferation of leukemic blasts in both the bone marrow and periphery (Döhner et al., 2022; Heuser et al., 2020).

The application of antibody-based TCEs in AML is hampered by the lack of TAAs with strongly reduced expression on healthy hematopoietic cells. Several TCEs targeting single antigens, such as CD33 or CD123, have failed to safely reach exposure levels in AML patients required for efficacy (Boyiadzis et al., 2023; R. Narayan et al., 2024). As a result, considerable attention has moved towards non-antibody based TCEs targeting multiple TAAs, as a means to gain selectivity towards AML cells (Zeng et al., 2024). MP0533 is a unique tetra-specific DARPin targeting three independent TAAs (CD33, CD123, and CD70) expressed by LSCs and leukemic blasts, and includes an affinity-tuned anti-CD3 DARPin domain for activation of T cells (Figure 2; (Bianchi et al., 2024)). This design aimed to selectively target AML cells and also to tackle the heterogeneous TAA expression common to AML. Two adjacent HSA-binding DARPin domains are included at the N-terminus of MP0533 to increase systemic half-life. Extensive preclinical studies were conducted to characterize the mechanism of action of MP0533, which confirmed the preferential activation of T cells in the presence of two or more TAAs on AML cells (Bianchi et al., 2024).

The first-in-human study in patients with r/r AML or myelodysplastic syndrome (MDS)/AML was initiated in early 2023 (NCT05673057). At the 66th American Society of Hematology Annual Meeting (December 7-10, 2024), interim data obtained from 37 patients treated with MP0533 with the first 7 escalating dose regimens were reported (Jongen-Lavrencic et al., 2024). A dosing scheme involving step-up dosing (SUD) followed by Q1W dosing at the target dose was devised with the aim of limiting the frequency and intensity of cytokine release syndrome (CRS); a common adverse event associated with CD3 engagers (Géraud et al., 2024). CRS (and related IRR) events predominantly occurred during treatment cycle 1 (28 days), typically within 24 h following MP0533 administration and two DLTs were reported at dose-level 7 (Jongen-Lavrencic et al., 2024). Clinical responses in 4 of the 37 patients at data cut-off (morphologically leukemia-free state, MLFS (n=3); complete remission, CR (n=1)) were observed across four intermediate dose regimens. In addition, 8 of 30 evaluable patients displayed ≥50% blast reduction in the bone marrow, including 6 of 13 patients with low diseases burden at baseline (<20% blasts), further supporting clinical activity of MP0533.

At the dose levels explored to date, free serum MP0533 exposure during the dosing interval was limited (Jongen-Lavrencic et al., 2024). The rapid clearance of free MP0533 may be due to two distinct mechanisms. Firstly, the strong avidity of MP0533, driven by simultaneous binding to three TAAs, likely results in pronounced TMDD. Secondly, ADA formation at the end of cycle 1 has been observed which may further limit exposure in later cycles. To date, no impact of ADAs on the safety profile of MP0533 has been observed.

Biomarker analyses both on bone marrow biopsies and peripheral blood demonstrated target engagement, evidenced by MP0533 detection on AML blasts, and modulation of sCD33 and IL-3 (CD123 ligand) levels (Jongen-Lavrencic et al., 2024). T cell activation in the periphery was demonstrated by increased expression of granzyme B+ and CD69+ on CD8+ T cells 24 h post-dose. Significantly, circulating levels of IFNγ and CXCL10, amongst other immune cell-derived mediators, were elevated in higher dose cohorts on cycle 1 day 15 (target dose) at 4h and 24h post-infusion, suggesting a potential dose-response relationship.

At the time of this publication, recruitment in the NCT05673057 phase 1 trial is ongoing. The acceptable safety profile and encouraging initial antitumor activity to-date supports further dose escalation. A clinical protocol amendment is in development for higher and more frequent dosing with the aim of improving MP0533 exposure and ultimately increasing response rate, depth, and durability, in the highly heterogeneous r/r AML population included in the study.

### Imaging with radiolabeled DARPins

Beyond the therapeutic applications of DARPins described above, dedicated efforts have been undertaken to assess the clinical utility of DARPins as target-specific radiolabeled tracers for diagnostic and imaging purposes. Such radiolabeled tracers ideally exhibit high specificity and affinity as well as fast clearance to achieve high imaging contrast in the first hours after administration. Depending on the radiolabel chosen, single photon emission computed tomography (SPECT)-based or positron emission tomography (PET)-based imaging is possible with differing advantages (Crișan et al., 2022).

The first phase 1 clinical imaging study with a DARPin was performed in patients with either HER2-positive or -negative (as defined by immunohistochemistry [IHC]) primary breast cancer (NCT04277338; (Bragina et al., 2022)), using a HER2-specific DARPin with picomolar affinity called G3 (Zahnd et al., 2007). A total of 28 patients underwent SPECT imaging, and three different doses (1, 2, or 3 mg) with about 300 MBq of ^99m^Tc as radioactive tracer were tested. Consistent with data from preclinical experiments, and the absence of half-life extension design features, ^99m^Tc-(HE)_3_-G3 was cleared rapidly from the circulation (elimination half-life of 3-4 h) and mostly accumulated in the kidney. No adverse effects were reported in patients administered with ^99m^Tc-(HE)_3_-G3. A significantly higher imaging contrast for ^99m^Tc-(HE)_3_-G3 was observed in patients demonstrating higher IHC-based HER2 expression. Furthermore, metastatic lesions in many organs including the bone and liver could be detected with ^99m^Tc-(HE)_3_-G3. Based on the data from this study, the authors recommended use of SPECT imaging with ^99m^Tc-(HE)_3_-G3 to distinguish HER2-positive and -negative breast cancer, including in metastatic lesions.

An additional phase 1 clinical study in patients with HER2 positive breast cancer was performed with ^99m^Tc-(HE)_3_-G3 (NCT05376644; (Bragina et al., 2023)). This study focused on direct in-patient comparison of the DARPin to another protein-based imaging agent called ADAPT6 (albumin-binding domain-derived affinity protein 6). While the G3 DARPin has about twice the molecular weight compared to ADAPT6 (14 kDa versus 5-7 kDa), the DARPin affinity for HER2 is higher (90 pM versus 1 nM). Both SPECT-imaging agents were reported to provide very good visualization properties. The ADAPT6 showed higher tumor accumulation whereas the DARPin showed higher contrast to background. Importantly, preclinical publications have shown that the G3 DARPin binds a distinct HER2 epitope compared to trastuzumab, as evidenced by the use of ^99m^Tc-(HE)_3_-G3 for monitoring of HER2 expression in xenografted human SKOV-3 ovarian cancer cells in mice during trastuzumab treatment (Jost et al., 2013; Tolmachev et al., 2022). Given the potential role of HER2 downmodulation as a mechanism of acquired resistance to anti-HER2 therapy (reviewed in (Wang et al., 2022)), ^99m^Tc-(HE)_3_-G3 could represent a valuable imaging tool for longitudinal and non-invasive monitoring of HER2 expression to guide the choice of HER2-targeted therapy.

A third DARPin imaging study was performed by the same research group in lung cancer patients using a DARPin targeting the epithelial cell adhesion molecule (EpCAM; NCT05620472; (Zelchan et al., 2024)). Three milligrams of the EpCAM-targeting DARPin (Ec1) were injected after labeling with ^99m^Tc at a clinical dose of about 300 MBq. Similar to the clinical experience with the HER2-targeting DARPin imaging agent described above, no adverse events were reported in patients administered ^99m^Tc-(HE)_3_-Ec1 in this single-dose study. SPECT-imaging confirmed that ^99m^Tc-(HE)_3_-Ec1 was effective for identification of EpCAM-expressing lung tumors and regional metastases. The kidney and liver were identified as the organs with highest accumulation of ^99m^Tc-(HE)_3_-Ec1. The authors concluded that this DARPin-based imaging agent has utility as a diagnostic for assessing EpCAM expression.

In conclusion, DARPin-based imaging agents have been successfully tested clinically and have shown their potential for diagnostic target-specific imaging of tumor lesions. As also reported with peptides and other mini-proteins, DARPins have demonstrated relatively high kidney accumulation and this observation has led to significant efforts in protein engineering to improve DARPin biodistribution (as discussed further below). Clinical studies are currently in planning to confirm the therapeutic potential of radio-labelled DARPins.

### Discussion and concluding perspectives for DARPin-based drug development – What did we learn? Where does it take us?

Over the last decade, clinical studies with DARPins have included >750 patients treated systemically across oncology and virology settings and have provided valuable insights into the utility of this novel protein drug class. The clinical experience to date has indicated that DARPins with both simple and highly sophisticated designs (e.g., multi-targeting, effector functions) can be readily generated, and that DARPins can be developed towards clinical application with exceptional speed (Rothenberger et al., 2022). This favorable ‘developability’ has been facilitated by the stable architecture of DARPins, as well as the ability to manufacture them in large quantities using microorganism-based expression systems, a rapid and cost-effective approach (Plückthun, 2015; Stumpp et al., 2020). DARPin drugs have proved suitable for systemic i.v. administration on diverse treatment regimens (i.e., single- or repeated-dosing), and the incorporation of half-life extension features has enabled tailoring of PK properties to the desired therapeutic modality. The safety profiles of DARPin drugs have typically been driven by the target biology (e.g., hypertension for VEGF-A inhibition) or the drug’s mechanism of action (e.g., CRS for TCEs), and no “class-specific” safety liabilities have been identified that would preclude the application of DARPins in specific patient populations.

Despite the many positives of the clinical experience with DARPins, immunogenicity has been a potential hindrance for some DARPins tested in clinical studies. Immunogenicity is a known phenomenon for protein-based therapeutics: monoclonal antibodies, Fc-fusion proteins, and camelid-derived nanobodies can also elicit immunogenicity to varying degrees, which can result in the generation of ADAs (Ackaert et al., 2021; Harris and Cohen, 2024). Diverse factors influence immunogenicity including the drug‘s mode of action (e.g., immune cell modulation), amino acid sequence, formulation, impurities, post-translational modifications (e.g., glycosylation), synthetic alterations (e.g., PEGylation), loss of structural integrity (e.g., aggregation, oxidation), and patient-specific factors (e.g., immune suppression) (Carter and Quarmby, 2024). ADAs can impact a drug’s pharmacokinetics (e.g. drug-sustaining or clearing) and biological activity (e.g. neutralizing) and in selected cases, ADAs have been associated with pronounced reduction of drug efficacy (Ridker et al., 2017) or safety (Carter and Quarmby, 2024; Chung et al., 2008). Therefore, the assessment of immunogenicity is an important requirement for the development of protein-based therapeutics (Shankar et al., 2014). As described above, ADAs against DARPins were reported in clinical studies, with varying impact on drug exposure (**Table 3**). In the cases of MP0250 (VEGF x HGF) and MP0317 (FAP x CD40), ADAs were detected in some patients, but these had minimal or no impact on drug exposure or biological activity. Conversely, ADAs against MP0310 (FAP x 4-1BB) and MP0533 (CD33 x CD123 x CD70 x CD3) were associated with reduced drug exposure. Thus, like for other therapeutic proteins, immunogenicity needs to be studied for each drug candidate and can affect them differently. Importantly, ADAs were not linked to major changes of the safety profile of DARPin drugs in any clinical studies conducted to date. Recent advances in computational prediction tools (Howard, Goens, Susta, Patel, and Wootton, 2025) and *in vitro* human immune cell assays (Karle, 2020) for the identification of putative T cell epitopes have opened the possibility to utilize sequence engineering to reduce the immunogenicity risk of candidate protein therapeutics. The robust stability and structural integrity of the DARPin scaffold makes this protein drug class highly amenable to sequence modification, and engineering for reduced immunogenicity potential is an ongoing and integral step in the development of the next generation of DARPin candidates. Additionally, a third-generation DARPin library lacking putative T cell epitopes (as evidenced by low MHC binding, reduced peptide presentation, and low inherent T cell stimulation in the appropriate pre-clinical assays) has been generated to accelerate the discovery of DARPin drug candidates with low immunogenicity risk (Reichen, 2025). Furthermore, recent advances in computer-aided protein design are being applied to design proteins (Jiang et al., 2024) and can be actively utilized in DARPin drug discovery efforts. These approaches have the potential to accelerate the generation of DARPin candidates and ultimately expedite their translation to clinical settings where they can benefit patients in need.

**Table 3.** Overview of clinical characteristics of systemically administered therapeutic DARPins.

| **DARPin** | **Antitumor efficacy** | **Monotherapy**<br>**MTD** | **Serum**<br>**half-life** | **Impact of ADAs**<br>**on exposure** | **Pharmacodynamic**<br>**effects observed** | **References** |
| --- | --- | --- | --- | --- | --- | --- |
| MP0250<br>*solid tumors* | 1/40 uPR, 24/40 SD | 8 mg/kg Q2W or<br>12 mg/kg Q3W | 11-16 days | None | VEGF-A↓, HGF↑ | (Baird et al., 2020) |
| MP0250<br>*MM\** | 9/28 PR, 3/28 MR, 9/28 SD | 8 mg/kg Q3W | 11-15 days | None | VEGF-A↓, HGF↑ | (Knop et al., 2024) |
| MP0274 | 1/22 PR, 3/22 SD | Not reached | 15 days | Not reported | Not reported | (Fischer et al., 2022) |
| MP0310 | 1/37 uPR, 16/37 SD | Not reached | Not<br>reported | Reduced<br>exposure | Not reported | Manuscript in<br>preparation |
| MP0317 | 1/46 uPR, 11/46 SD | Not reached | Not<br>reported | None at higher<br>doses | sFAP↓, sCD40↑,<br>TME remodeling,<br>CXCL10↑ | (Krieg et al., 2024;<br>Steeghs et al., 2024) |
| MP0420 /<br>*ensovibep* | N.A. | Not reached | ~15 days | None during<br>acute viral<br>infection phase | Viral titer↓ | (Kingsley et al., 2024;<br>Prins et al., 2022;<br>Stojcheva et al., 2023) |
| MP0533<sup>#</sup> | 1/30 CR, 3/30 MLFS<sup>#</sup> | Not yet defined<sup>#</sup> | To be<br>determined<sup>#</sup> | Reduced<br>exposure | IL-3↑, sCD33↓, T<br>cell activation | (Jongen-Lavrencic et<br>al., 2024) |

\* Combination with bortezomib and dexamethasone. # Study ongoing
Abbreviations: ADA, anti-drug antibody; CR, complete remission; MLFS, morphologic leukemia-free state; MM, multiple myeloma; MR, minimal response; MTD, maximally tolerated dose; N.A., not applicable; PR, partial response; sCD33, soluble CD33; sCD40, soluble CD40; sFAP, soluble FAP; SD, stable disease; uPR, unconfirmed partial response; Q2W, dosing once every two weeks; Q3W, dosing once every three weeks; TME, tumor microenvironment.

The last decade has seen the evolution from relatively ‘simple’ growth factor inhibitor DARPins (e.g., abicipar) to more sophisticated multi-specific DARPins (e.g., MP0274), and DARPins that modulate the activity of immune effector cells (e.g., MP0317). This transition is poised to continue with a new generation of logic-gated multi-specific immune cell engagers called “Switch-DARPins”. These DARPin candidates contain a 2-in-1 DARPin (the Switch) which has the possibility to bind to two different targets in a mutually exclusive way (2-in-1 or “either-or” Switch-DARPin) and allows conditional activation/binding of either one or the other target depending on the biological context/environment. Recently published preclinical data has given the first insights into the potential of Switch-DARPins for conditional and reversible activation of target cell killing by NK cells (Link et al., 2024) or T cells (Robinson et al., 2024). These early findings provide an impetus for further investigation of the potential of Switch-DARPins, particularly in disease settings that are inaccessible to antibody-based immune engaging drugs.

Considerable attention has recently also been directed towards the potential application of DARPins as vectors/ligands for targeted delivery of radioactive payloads to cancer cells for diagnostic and therapeutic purposes. Early clinical imaging studies with radio-labelled DARPins binding HER2 and EpCAM have confirmed selective uptake at tumor lesions, but have also suggested that optimization of biodistribution properties, in particular kidney accumulation, is of key interest for DARPin-based radioligand therapy. The ^212^Pb-labelled Radio-DARPin candidate MP0712, which targets the delta-like ligand 3 (DLL3) antigen expressed on small cell lung cancer (SCLC) cells and other neuroendocrine tumors (NETs), has an attractive preclinical biodistribution profile (reaching a tumor to kidney ratio > 2 within the first 24 hours) that enabled anti-tumor efficacy and favorable safety in mice (Croset et al., 2024; Lizak et al., 2024). Engineering of DARPins to reduce kidney accumulation, alongside high affinity DLL3 binding and half-life extension properties, were described as critical elements in the development of MP0712, which should also be applied to other Radio-DARPins. Clinical examination of the therapeutic profiles of Radio-DARPins, like MP0712, will be of considerable interest in the future and of high potential value for SCLC patients.

In summary, DARPins have been developed to date through a range of clinical phases (up to pivotal stage) and in diverse indications and settings, including in healthy volunteers, radio-imaging studies, and both monotherapy and combination therapy trials. Several of these studies have demonstrated that DARPins can deliver meaningful clinical efficacy to patients, consistent with the drug’s intended mechanism of action. DARPins continue to demonstrate their versatility for increasingly sophisticated drug architectures, in some cases enabling disease-targeting therapeutic modalities that were previously inaccessible to other protein-based drug classes (e.g. MP0533 with avidity-gated AML cell killing). Building on these clinical data and further promising preclinical findings, the next generations of novel DARPin-based drugs, including Radio- and Switch-DARPins, are planned to undergo clinical examination in the near future. These studies will ultimately reveal the therapeutic potential of the novel DARPin-based candidates for the treatment of cancer indications with high unmet need.

## Data Availability

This systematic review summarized work published to date. Please see reviewed research for data availability.

## Acknowledgements

This systematic review was supported by Molecular Partners AG. All DARPin therapeutic studies except ACTIV-3 were sponsored by Molecular Partners AG. The ACTIV-3 study was partially funded by the National Institutes of Health.

The Authors would like to thank all patients, care givers, and investigators as well as the team members at the collaboration partners Amgen and Novartis, who made those studies possible. Marianne Jenal-Eyholzer, PhD, CMPP at Molecular Partners AG is gratefully acknowledged for support with the systematic literature search and critical review of the manuscript.

## Review registration and protocol

This systematic review was not registered and a protocol was not prepared. The search strategy is described in detail in the methods and results are reported descriptively. No meta-analysis or other synthesis of the results was performed.

## Ethic approvals and compliance with GCP

Requirements for ethical and regulatory approvals as well as compliance with the declaration of Helsinki and the guidelines for ICH GCP, including written informed consent of all patients prior to study start, are described in the respective primary publications of the studies summarized in this review.

## References

Ackaert, C., Smiejkowska, N., Xavier, C., Sterckx, Y. G. J., Denies, S., Stijlemans, B., … Keyaerts, M. (2021). Immunogenicity Risk Profile of Nanobodies. Frontiers in Immunology, 12, 632687. 10.3389/fimmu.2021.632687

ACTIV-3/TICO_Bamlanivimab_Study_Group*, Lundgren, J. D., Grund, B., Barkauskas, C. E., Holland, T. L., Gottlieb, R. L., … Wenner, C. (2022). Responses to a Neutralizing Monoclonal Antibody for Hospitalized Patients With COVID-19 According to Baseline Antibody and Antigen Levels: A Randomized Controlled Trial. Annals of Internal Medicine, 175(2), 234–243. 10.7326/m21-3507

ACTIV-3/TICO_LY-CoV555_Study_Group, Lundgren, J. D., Grund, B., Barkauskas, C. E., Holland, T. L., Gottlieb, R. L., … Neaton, J. D. (2020). A Neutralizing Monoclonal Antibody for Hospitalized Patients with Covid-19. New England Journal of Medicine, 384(10), 905– 914. 10.1056/nejmoa2033130

ACTIV-3/TICO_Study_Group, Barkauskas, C., Mylonakis, E., Poulakou, G., Young, B. E., Vock, D. M., … Williams, R. (2022). Efficacy and Safety of Ensovibep for Adults Hospitalized With COVID-19. Annals of Internal Medicine, 175(9), M22–1503. 10.7326/m22-1503

ACTIV-3/TICO_Study_Group, Self, W. H., Sandkovsky, U., Reilly, C. S., Vock, D. M., Gottlieb, R. L., … Wenner, C. (2022). Efficacy and safety of two neutralising monoclonal antibody therapies, sotrovimab and BRII-196 plus BRII-198, for adults hospitalised with COVID-19 (TICO): a randomised controlled trial. The Lancet Infectious Diseases, 22(5), 622–635. 10.1016/s1473-3099(21)00751-9

Aerts, J., Paz-Ares, L. G., Helissey, C., Cappuzzo, F., Quere, G., Kowalski, D., … Felip, E. (2024). Acasunlimab (DuoBody-PD-L1×4-1BB) alone or in combination with pembrolizumab (pembro) in patients (pts) with previously treated metastatic non-small cell lung cancer (mNSCLC): Initial results of a randomized, open-label, phase 2 trial. Journal of Clinical Oncology, 42(16_suppl), 2533–2533. 10.1200/jco.2024.42.16_suppl.2533

Anderson, W. J., da Cruz, N. F. S., Lima, L. H., Emerson, G. G., Rodrigues, E. B., and Melo, G. B. (2021). Mechanisms of sterile inflammation after intravitreal injection of antiangiogenic drugs: a narrative review. International Journal of Retina and Vitreous, 7(1), 37. 10.1186/s40942-021-00307-7

Baird, R. D., Linossi, C., Middleton, M., Lord, S., Harris, A., Rodón, J., … Omlin, A. (2020). First-in-Human Phase I Study of MP0250, a First-in-Class DARPin Drug Candidate Targeting VEGF and HGF, in Patients With Advanced Solid Tumors. Journal of Clinical Oncology, 39(2), 145–154. 10.1200/jco.20.00596

Bianchi, M., Reichen, C., Croset, A., Fischer, S., Eggenschwiler, A., Grübler, Y., … Goubier, A. (2024). The CD33xCD123xCD70 Multispecific CD3-Engaging DARPin MP0533 Induces Selective T Cell–Mediated Killing of AML Leukemic Stem Cells. Cancer Immunology Research, OF1–OF23. 10.1158/2326-6066.cir-23-0692

Binz, H. K., Bakker, T. R., Phillips, D. J., Cornelius, A., Zitt, C., Göttler, T., … Stumpp, M. T. (2017). Design and characterization of MP0250, a tri-specific anti-HGF/anti-VEGF DARPin® drug candidate. MAbs, 9(8), 1262–1269. 10.1080/19420862.2017.1305529

Boyiadzis, M., Desai, P., Daskalakis, N., Donnellan, W., Ferrante, L., Goldberg, J. D., … Alonso-Dominguez, J. M. (2023). First-in-human study of JNJ-63709178, a CD123/CD3 targeting antibody, in relapsed/refractory acute myeloid leukemia. Clinical and Translational Science, 16(3), 429–435. 10.1111/cts.13467

Bragina, O., Chernov, V., Schulga, A., Konovalova, E., Garbukov, E., Vorobyeva, A., … Tolmachev, V. (2022). Phase I Trial of 99mTc-(HE)3-G3, a DARPin-Based Probe for Imaging of HER2 Expression in Breast Cancer. Journal of Nuclear Medicine, 63(4), 528–535. 10.2967/jnumed.121.262542

Bragina, O., Chernov, V., Schulga, A., Konovalova, E., Hober, S., Deyev, S., … Tolmachev, V. (2023). Direct Intra-Patient Comparison of Scaffold Protein-Based Tracers, [99mTc]Tc-ADAPT6 and [99mTc]Tc-(HE)3-G3, for Imaging of HER2-Positive Breast Cancer. Cancers, 15(12), 3149. 10.3390/cancers15123149

Carter, P. J., and Quarmby, V. (2024). Immunogenicity risk assessment and mitigation for engineered antibody and protein therapeutics. Nature Reviews Drug Discovery, 1–16. 10.1038/s41573-024-01051-x

Chester, C., Sanmamed, M. F., Wang, J., and Melero, I. (2018). Immunotherapy targeting 4-1BB: mechanistic rationale, clinical results, and future strategies. Blood, 131(1), 49–57. 10.1182/blood-2017-06-741041

Chung, C. H., Mirakhur, B., Chan, E., Quynh-Thu, L., Jordan, B., Michael, M., … Platts-Mills, T. A. E. (2008). Cetuximab-Induced Anaphylaxis and IgE Specific for Galactose-α-1,3-Galactose. New England Journal of Medicine, 358(11), 1109–1117. 10.1056/nejmoa074943

Claas, A. M., Lee, M., Huang, P., Knutson, C. G., Bullara, D., Schoeberl, B., and Gaudet, S. (2024). Viral Kinetics Model of SARS-CoV-2 Infection Informs Drug Discovery, Clinical Dose, and Regimen Selection. Clinical Pharmacology & Therapeutics, 116(3), 757–769. 10.1002/cpt.3267

Claus, C., Ferrara-Koller, C., and Klein, C. (2023). The emerging landscape of novel 4-1BB (CD137) agonistic drugs for cancer immunotherapy. MAbs, 15(1), 2167189. 10.1080/19420862.2023.2167189

Cortot, A., Le, X., Smit, E., Viteri, S., Kato, T., Sakai, H., … Paik, P. K. (2022). Safety of MET Tyrosine Kinase Inhibitors in Patients With MET Exon 14 Skipping Non-small Cell Lung Cancer: A Clinical Review. Clinical Lung Cancer, 23(3), 195–207. 10.1016/j.cllc.2022.01.003

Crișan, G., Moldovean-Cioroianu, N. S., Timaru, D.-G., Andrieș, G., Căinap, C., and Chiș, V. (2022). Radiopharmaceuticals for PET and SPECT Imaging: A Literature Review over the Last Decade. International Journal of Molecular Sciences, 23(9), 5023. 10.3390/ijms23095023

Croset, A., Saidi, A., Malvezzi, F., Lizak, C., Torgue, J., and Steiner, D. (2024). EANM’24 Abstract Book Congress Oct 19-23, 2024. European Journal of Nuclear Medicine and Molecular Imaging, 51(Suppl 1), 1–1026. 10.1007/s00259-024-06838-z

Desideri, L. F., Traverso, C. E., and Nicolò, M. (2020). Abicipar pegol: an investigational anti-VEGF agent for the treatment of wet age-related macular degeneration. Expert Opinion on Investigational Drugs, 29(7), 651–658. 10.1080/13543784.2020.1772754

Döhner, H., Wei, A. H., Appelbaum, F. R., Craddock, C., DiNardo, C. D., Dombret, H., … Löwenberg, B. (2022). Diagnosis and management of AML in adults: 2022 recommendations from an international expert panel on behalf of the ELN. Blood, 140(12), 1345–1377. 10.1182/blood.2022016867

Dolado, I., Fiedler, U., Strobel, H., Metz, C., Stumpp, M., and Rojkjaer, L. (2013). Abstract P4-12-30: A bivalent Her2 targeting DARPin with high efficacy against Her2-low and Her2-positive tumors. Cancer Research, 73(24_Supplement), P4-12-30-P4-12–30. 10.1158/0008-5472.sabcs13-p4-12-30

Dziadek, S., Kraxner, A., Cheng, W.-Y., Yang, T.-H. O., Flores, M., Theiss, N., … Charo, J. (2024). Comprehensive analysis of fibroblast activation protein expression across 23 tumor indications: insights for biomarker development in cancer immunotherapies. Frontiers in Immunology, 15, 1352615. 10.3389/fimmu.2024.1352615

Ferrucci, A., Moschetta, M., Frassanito, M. A., Berardi, S., Catacchio, I., Ria, R., … Vacca, A. (2014). A HGF/cMET Autocrine Loop Is Operative in Multiple Myeloma Bone Marrow Endothelial Cells and May Represent a Novel Therapeutic Target. Clinical Cancer Research, 20(22), 5796–5807. 10.1158/1078-0432.ccr-14-0847

Fiedler, U., Ekawardhani, S., Cornelius, A., Gilboy, P., Bakker, T. R., Dolado, I., … Dawson, K. M. (2017). MP0250, a VEGF and HGF neutralizing DARPin® molecule shows high anti-tumor efficacy in mouse xenograft and patient-derived tumor models. Oncotarget, 8(58), 98371–98383. 10.18632/oncotarget.21738

Fiedler, U., Metz, C., Zitt, C., Bessey, R., Béhé, M., Blanc, A., … Kiemle-Kallee, J. (2017). Abstract P4-21-18: Pre-clinical antitumor activity, tumor localization, and pharmacokinetics of MP0274, an apoptosis inducing, biparatopic HER2-targeting DARPin®. Cancer Research, 77(4_Supplement), P4-21-18-P4-21–18. 10.1158/1538-7445.sabcs16-p4-21-18

Fischer, S., Götze, T. O., Omlin, A., Baird, R. D., Dawson, K. M., Zitt, C., … Fremd, C. (2022). A Case of Sustained Tumor Regression With MP0274, a Novel DARPin Therapeutic Targeting Human Epidermal Growth Factor Receptor 2 Signaling, in Metastatic Human Epidermal Growth Factor Receptor 2–Positive Breast Cancer After Prior Trastuzumab and Pertuzumab. JCO Precision Oncology, 6(6), e2200006. 10.1200/po.22.00006

Géraud, A., Hueso, T., Laparra, A., Bige, N., Ouali, K., Cauquil, C., … Michot, J.-M. (2024). Reactions and adverse events induced by T-cell engagers as anti-cancer immunotherapies, a comprehensive review. European Journal of Cancer, 205, 114075. 10.1016/j.ejca.2024.114075

Harris, C. T., and Cohen, S. (2024). Reducing Immunogenicity by Design: Approaches to Minimize Immunogenicity of Monoclonal Antibodies. BioDrugs, 38(2), 1–22. 10.1007/s40259-023-00641-2

Heuser, M., Ofran, Y., Boissel, N., Mauri, S. B., Craddock, C., Janssen, J., … Committee, E. G. (2020). Acute myeloid leukaemia in adult patients: ESMO Clinical Practice Guidelines for diagnosis, treatment and follow-up † † Approved by the ESMO Guidelines Committee: August 2002, last update January 2020. This publication supersedes the previously published version—Ann Oncol. 2013;24(suppl 6):vi138–vi143. *Annals of Oncology*, *31*(6), 697–712. 10.1016/j.annonc.2020.02.018

Holland, T. L., Ginde, A. A., Paredes, R., Murray, T. A., Engen, N., Grandits, G., … Lundgren, J. D. (2022). Tixagevimab–cilgavimab for treatment of patients hospitalised with COVID-19: a randomised, double-blind, phase 3 trial. The Lancet Respiratory Medicine, 10(10), 972–984. 10.1016/s2213-2600(22)00215-6

Howard, E. L., Goens, M. M., Susta, L., Patel, A., and Wootton, S. K. (2025). Anti-Drug Antibody Response to Therapeutic Antibodies and Potential Mitigation Strategies. Biomedicines, 13(2), 299. 10.3390/biomedicines13020299

Jian, C.-Z., Lin, L., Hsu, C.-L., Chen, Y.-H., Hsu, C., Tan, C.-T., and Ou, D.-L. (2024). A potential novel cancer immunotherapy: Agonistic anti-CD40 antibodies. Drug Discovery Today, 29(3), 103893. 10.1016/j.drudis.2024.103893

Jiang, H., Jude, K. M., Wu, K., Fallas, J., Ueda, G., Brunette, T. J., … Baker, D. (2024). De novo design of buttressed loops for sculpting protein functions. Nature Chemical Biology, 1–7. 10.1038/s41589-024-01632-2

Jongen-Lavrencic, M., Pabst, T., Bories, P., Griškevičius, L., Huls, G., de Leeuw, D. C., … Subklewe, M. (2024). MP0533 (CD33 x CD123 x CD70 x CD3), a Tetra-Specific CD3-Engaging Darpin for the Treatment of Patients with Relapsed/Refractory AML or MDS/AML: Results of an Ongoing Phase 1/2a Study. Blood, 144(Supplement 1), 2881. 10.1182/blood-2024-199211

Jost, C., Schilling, J., Tamaskovic, R., Schwill, M., Honegger, A., and Plückthun, A. (2013). Structural Basis for Eliciting a Cytotoxic Effect in HER2-Overexpressing Cancer Cells via Binding to the Extracellular Domain of HER2. Structure, 21(11), 1979–1991. 10.1016/j.str.2013.08.020

Karle, A. C. (2020). Applying MAPPs Assays to Assess Drug Immunogenicity. Frontiers in Immunology, 11, 698. 10.3389/fimmu.2020.00698

Khurana, R. N., Kunimoto, D., Yoon, Y. H., Wykoff, C. C., Chang, A., Maturi, R. K., … CEDAR_and_SEQUOIA_Study_Groups. (2021). Two-Year Results of the Phase 3 Randomized Controlled Study of Abicipar in Neovascular Age-Related Macular Degeneration. Ophthalmology, 128(7), 1027–1038. 10.1016/j.ophtha.2020.11.017

Kingsley, J., Kumarasamy, N., Abrishamian, L., Bonten, M., Igbinadolor, A., Mekebeb-Reuter, M., … Chandra, R. (2024). The DARPin antiviral ensovibep for non-hospitalized patients with COVID-19: Results from EMPATHY, a randomized, placebo-controlled Phase 2 study. Open Forum Infectious Diseases, 11(6), ofae233. 10.1093/ofid/ofae233

Knop, S., Szarejko, M., Grząśko, N., Bringhen, S., Trautmann-Grill, K., Jurczyszyn, A., … Goldschmidt, H. (2024). A phase 1b/2 study evaluating efficacy and safety of MP0250, a designed ankyrin repeat protein (DARPin) simultaneously targeting vascular endothelial growth factor (VEGF) and hepatocyte growth factor (HGF), in combination with bortezomib and dexamethasone, in patients with relapsed or refractory multiple myeloma. EJHaem. 10.1002/jha2.968

Krieg, J., Stavropoulou, V., Ribeiro, A., Florescu, A.-M., Winter, H. D., Stojcheva, N., … Legenne, P. (2024). 612 Comprehensive biomarker analyses from a phase 1 study reveals marked tumor microenvironment modulation in patients with advanced solid tumors treated with MP0317, a FAP-localized CD40 agonistic DARPin. Regular and Young Investigator Award Abstracts, A704–A704. 10.1136/jitc-2024-sitc2024.0612

Kunimoto, D., Yoon, Y. H., Wykoff, C. C., Chang, A., Khurana, R. N., Maturi, R. K., … CEDAR_and_SEQUOIA_Study_Groups. (2020). Efficacy and Safety of Abicipar in Neovascular Age-Related Macular Degeneration 52-Week Results of Phase 3 Randomized Controlled Study. Ophthalmology, 127(10), 1331–1344. 10.1016/j.ophtha.2020.03.035

Lin, C. H.-T., Tariq, M. J., Ullah, F., Sannareddy, A., Khalid, F., Abbas, H., … Dima, D. (2024). Current Novel Targeted Therapeutic Strategies in Multiple Myeloma. International Journal of Molecular Sciences, 25(11), 6192. 10.3390/ijms25116192

Link, A., Frasconi, T. M., Wullschleger, S., Venetz, N., Ribeiro, A., Schlegel, A., … Goubier, A. (2024). MP0621 (cKit x CD16a x CD47), a Multi-Specific Switch-Darpin with Conditional Blockade of CD47 Targeting Hematopoietic Stem Cells: Preclinical Evaluation of a Next-Generation Conditioning Agent for Stem Cell Transplantation. Blood, 144(Supplement 1), 4775–4775. 10.1182/blood-2024-199982

Link, A., Hepp, J., Reichen, C., Schildknecht, P., Tosevski, I., Taylor, J., … vom Baur, E. (2018). Abstract 3752: Preclinical pharmacology of MP0310: A 4-1BB/FAP bispecific DARPin drug candidate promoting tumor-restricted T-cell costimulation. Cancer Research, 78(13_Supplement), 3752–3752. 10.1158/1538-7445.am2018-3752

Link, A., Juglair, L., Poulet, H., Lemaillet, G., Reichen, C., Schildknecht, P., … Baur, E. V. (2020). Abstract 2273: Selection of first-in-human clinical dose range for the tumor-targeted 4-1BB agonist MP0310 (AMG 506) using a pharmacokinetic/pharmacodynamics modeling approach. Cancer Research, 80(16_Supplement), 2273–2273. 10.1158/1538-7445.am2020-2273

Lizak, C., Malvezzi, F., Saidi, A., Mettier, M., Vojackova, J., Schibli, R., … Steiner, D. (2024). Lead-212 Radio-DARPin Therapeutic (RDT) targeting delta-like ligand 3 (DLL3) shows promising preclinical antitumor efficacy and tolerability in small cell lung cancer (SCLC). Journal of Nuclear Medicine June 2024, 65 *(supplement 2)*, 241995. Soc Nuclear Med.

Melero, I., Reis, B., Bardaji, M. J. L., Spanggaard, I., Lee, D. H., Spicer, J., … Epp, A. (2024). 161P Fibroblast activation protein (FAP)-CD40 (RO7300490) mediates intratumoral DC maturation and modulation of the tumor microenvironment. Annals of Oncology, 35, S279– S280. 10.1016/j.annonc.2024.08.169

Melo, G., Cruz, Emerson, Rezende, Meyer, Uchiyama, … Rodrigues, E. (2021). Critical analysis of techniques and materials used in devices, syringes, and needles used for intravitreal injections. Progress in Retinal and Eye Research, 80, 100862. 10.1016/j.preteyeres.2020.100862

Mitra, V., Fiedler, U., Snell, D., Dawson, K. M., Jung, S., Pike, I., and vom Baur, E. (2017). Abstract 4966: Phospho-proteome analyses confirm the unique mode of action of MP0274, an apoptosis inducing, biparatopic HER2-targeting DARPin® drug candidate. Cancer Research, 77(13_Supplement), 4966–4966. 10.1158/1538-7445.am2017-4966

Narayan, P., Osgood, C. L., Singh, H., Chiu, H.-J., Ricks, T. K., Chow, E. C. Y., … Beaver, J. A. (2020). FDA Approval Summary: Fam-Trastuzumab Deruxtecan-Nxki for the Treatment of Unresectable or Metastatic HER2-Positive Breast Cancer. Clinical Cancer Research: An Official Journal of the American Association for Cancer Research, 27(16), 4478–4485. 10.1158/1078-0432.ccr-20-4557

Narayan, R., Piérola, A. A., Donnellan, W. B., Yordi, A. M., Abdul-Hay, M., Platzbecker, U., … Esteban, D. (2024). First-in-human study of JNJ-67571244, a CD33 × CD3 bispecific antibody, in relapsed/refractory acute myeloid leukemia and myelodysplastic syndrome. Clinical and Translational Science, 17(3), e13742. 10.1111/cts.13742

Papadimitrakopoulou, V. A., Wu, Y.-L., Han, J.-Y., Ahn, M.-J., Ramalingam, S. S., John, T., … Mok, T. S. K. (2018). LBA51 Analysis of resistance mechanisms to osimertinib in patients with EGFR T790M advanced NSCLC from the AURA3 study. Annals of Oncology, 29, viii741. 10.1093/annonc/mdy424.064

Piha-Paul, S., Olwill, S. A., Hamilton, E., Tolcher, A., Pohlmann, P., Liu, S. V., … Ku, G. (2024). A First-in-Human Study of cinrebafusp alfa, a HER2/4-1BB Bispecific Molecule, in Patients with HER2-Positive Advanced Solid Malignancies. Clinical Cancer Research, OF1– OF11. 10.1158/1078-0432.ccr-24-1552

Plückthun, A. (2015). Designed Ankyrin Repeat Proteins (DARPins): Binding Proteins for Research, Diagnostics, and Therapy. Annual Review of Pharmacology and Toxicology, 55(1), 489–511. 10.1146/annurev-pharmtox-010611-134654

Prins, M. L. M., van der Plas, J. L., Vissers, M. F. J. M., Berends, C. L., Tresch, G., Soergel, M., … Kamerling, I. M. C. (2022). Viral clearance, pharmacokinetics and tolerability of ensovibep in patients with mild to moderate COVID-19: A phase 2a, open-label, single-dose escalation study. British Journal of Clinical Pharmacology, 89(3), 1105–1114. 10.1111/bcp.15560

Rao, L., Veirman, K. D., Giannico, D., Saltarella, I., Desantis, V., Frassanito, M. A., … Vacca, A. (2018). Targeting angiogenesis in multiple myeloma by the VEGF and HGF blocking DARPin® protein MP0250: a preclinical study. Oncotarget, 9(17), 13366–13381. 10.18632/oncotarget.24351

Reichen, C. (2025, May 13). DARPins for Radiotherapy. Presented at the Protein Engineering Global Summit (PEGS) 2025. Retrieved from https://www.pegsummit.com/display-of-biologics#ChristianReichen

Ridker, P. M., Tardif, J.-C., Amarenco, P., Duggan, W., Glynn, R. J., Jukema, J. W., … Investigators, S. (2017). Lipid-Reduction Variability and Antidrug-Antibody Formation with Bococizumab. The New England Journal of Medicine, 376(16), 1517–1526. 10.1056/nejmoa1614062

Rigamonti, N., Veitonmäki, N., Domke, C., Barsin, S., Jetzer, S., Abdelmotaleb, O., … Trail, P. A. (2022). A Multispecific Anti-CD40 DARPin Construct Induces Tumor-Selective CD40 Activation and Tumor Regression. Cancer Immunology Research, 10(5), 626–640. 10.1158/2326-6066.cir-21-0553

Robinson, J., Bianchi, M., Müller, M. R., Ems, G., Friang, C., Grübler, Y., … Ayala, M. G. (2024). 842 Unlocking precision: a next-generation multi-specific CD3 Switch-DARPin with enhanced function to tackle the current limitations of T cell engagers in ovarian cancer. *Regular and Young Investigator Award Abstracts*, A951–A951. 10.1136/jitc-2024-sitc2024.0842

Rothenberger, S., Hurdiss, D. L., Walser, M., Malvezzi, F., Mayor, J., Ryter, S., … Trimpert, J. (2022). The trispecific DARPin ensovibep inhibits diverse SARS-CoV-2 variants. Nature Biotechnology, 40(12), 1845–1854. 10.1038/s41587-022-01382-3

Saltarella, I., Altamura, C., Campanale, C., Laghetti, P., Vacca, A., Frassanito, M. A., and Desaphy, J.-F. (2023). Anti-Angiogenic Activity of Drugs in Multiple Myeloma. Cancers, 15(7), 1990. 10.3390/cancers15071990

Saltarella, I., Link, A., Lamanuzzi, A., Reichen, C., Robinson, J., Altamura, C., … Desaphy, J.-F. (2024). Improvement of daratumumab- or elotuzumab-mediated NK cell activity by the bi-specific 4-1BB agonist, DARPin α-FAPx4–1BB: A preclinical study in multiple myeloma. Biomedicine & Pharmacotherapy, 176, 116877. 10.1016/j.biopha.2024.116877

Segal, N. H., He, A. R., Doi, T., Levy, R., Bhatia, S., Pishvaian, M. J., … Gopal, A. K. (2018). Phase I Study of Single-Agent Utomilumab (PF-05082566), a 4-1BB/CD137 Agonist, in Patients with Advanced Cancer. Clinical Cancer Research, 24(8), 1816–1823. 10.1158/1078-0432.ccr-17-1922

Shankar, G., Arkin, S., Cocea, L., Devanarayan, V., Kirshner, S., Kromminga, A., … Scientists, A. A. of P. (2014). Assessment and Reporting of the Clinical Immunogenicity of Therapeutic Proteins and Peptides—Harmonized Terminology and Tactical Recommendations. The AAPS Journal, 16(4), 658–673. 10.1208/s12248-014-9599-2

Shouse, G. (2024). Bispecific antibodies for the treatment of hematologic malignancies: The magic is T-cell redirection. Blood Reviews, 101251. 10.1016/j.blre.2024.101251

Singh, R., Kim, Y.-H., Lee, S.-J., Eom, H.-S., and Choi, B. K. (2024). 4-1BB immunotherapy: advances and hurdles. Experimental & Molecular Medicine, 56(1), 32–39. 10.1038/s12276-023-01136-4

Stebbing, J., Copson, E., and O’Reilly, S. (2000). Herceptin (trastuzamab) in advanced breast cancer. Cancer Treatment Reviews, 26(4), 287–290. 10.1053/ctrv.2000.0182

Steeghs, N., Gomez-Roca, C. A., Korakis, I., Gort, E. H., Winter, H. A. M. D., Stojcheva, N., … Cassier, P. A. (2024). Effect of MP0317, a FAP x CD40 DARPin, on safety profile and tumor-localized CD40 activation in a phase 1 study in patients with advanced solid tumors. Journal of Clinical Oncology, 42(16_suppl), 2573–2573. 10.1200/jco.2024.42.16_suppl.2573

Steiner, D., Merz, F. W., Sonderegger, I., Gulotti-Georgieva, M., Villemagne, D., Phillips, D. J., … Binz, H. K. (2017). Half-life extension using serum albumin-binding DARPin® domains. *Protein Engineering*, Design & Selection, 30(9), 583–591. 10.1093/protein/gzx022

Stojcheva, N., Gladman, S., Soergel, M., Zitt, C., Drake, R., Lockett, T., … Boyce, M. (2023). Ensovibep, a SARS-CoV-2 antiviral designed ankyrin repeat protein, is safe and well tolerated in healthy volunteers: Results of a first-in-human, ascending single-dose Phase 1 study. British Journal of Clinical Pharmacology, 89(7), 2295–2303. 10.1111/bcp.15747

Stumpp, M. T., Dawson, K. M., and Binz, H. K. (2020). Beyond Antibodies: The DARPin® Drug Platform. BioDrugs, 34(4), 423–433. 10.1007/s40259-020-00429-8

Timmerman, J., Herbaux, C., Ribrag, V., Zelenetz, A. D., Houot, R., Neelapu, S. S., … Levy, R. (2020). Urelumab alone or in combination with rituximab in patients with relapsed or refractory B-cell lymphoma. American Journal of Hematology, 95(5), 510–520. 10.1002/ajh.25757

Tolmachev, V., Bodenko, V., Oroujeni, M., Deyev, S., Konovalova, E., Schulga, A., … Vorobyeva, A. (2022). Direct In Vivo Comparison of 99mTc-Labeled Scaffold Proteins, DARPin G3 and ADAPT6, for Visualization of HER2 Expression and Monitoring of Early Response for Trastuzumab Therapy. International Journal of Molecular Sciences, 23(23), 15181. 10.3390/ijms232315181

Vos, E. R. A., van Hagen, C. C. E., Wong, D., Smits, G., Kuijer, M., Wijmenga-Monsuur, A. J., … de Melker, H. E.. (2024). SARS-CoV-2 Seroprevalence Trends in the Netherlands in the Variant of Concern Era: Input for Future Response. Influenza and Other Respiratory Viruses, 18(6), e13312. 10.1111/irv.13312

Walser, M., Mayor, J., and Rothenberger, S. (2022). Designed Ankyrin Repeat Proteins: A New Class of Viral Entry Inhibitors. Viruses, 14(10), 2242. 10.3390/v14102242

Wang, Z., Zheng, Z., Jia, S., Liu, S., Xiao, X., Chen, G., … Lu, X. (2022). Trastuzumab resistance in HER2-positive breast cancer: Mechanisms, emerging biomarkers and targeting agents. Frontiers in Oncology, 12, 1006429. 10.3389/fonc.2022.1006429

Weinreich, D. M., Sivapalasingam, S., Norton, T., Ali, S., Gao, H., Bhore, R., … Investigators, T. (2021). REGEN-COV Antibody Combination and Outcomes in Outpatients with Covid-19. New England Journal of Medicine, 385(23), e81. 10.1056/nejmoa2108163

Wu, Y.-L., Guarneri, V., Voon, P. J., Lim, B. K., Yang, J.-J., Wislez, M., … investigators, I. 2. (2024). Tepotinib plus osimertinib in patients with EGFR-mutated non-small-cell lung cancer with MET amplification following progression on first-line osimertinib (INSIGHT 2): a multicentre, open-label, phase 2 trial. The Lancet Oncology, 25(8), 989–1002. 10.1016/s1470-2045(24)00270-5

Yewdell, J. W. (2021). Antigenic drift: Understanding COVID-19. Immunity, 54(12), 2681– 2687. 10.1016/j.immuni.2021.11.016

Zahnd, C., Wyler, E., Schwenk, J. M., Steiner, D., Lawrence, M. C., McKern, N. M., … Plückthun, A. (2007). A Designed Ankyrin Repeat Protein Evolved to Picomolar Affinity to Her2. Journal of Molecular Biology, 369(4), 1015–1028. 10.1016/j.jmb.2007.03.028

Zelchan, R., Chernov, V., Medvedeva, A., Rybina, A., Bragina, O., Mishina, E., … Tolmachev, V. (2024). Phase I Clinical Evaluation of Designed Ankyrin Repeat Protein [99mTc]Tc(CO)3-(HE)3-Ec1 for Visualization of EpCAM-Expressing Lung Cancer. Cancers, 16(16), 2815. 10.3390/cancers16162815

Zeng, Z., Roobrouck, A., Deschamps, G., Bonnevaux, H., Guerif, S., Brabandere, V. D., … Dullaers, M. (2024). Dual-targeting CD33/CD123 NANOBODY® T cell engager with potent anti-AML activity and good safety profile. Blood Advances. 10.1182/bloodadvances.2023011858

Zhang, Y.-W., Su, Y., Volpert, O. V., and Woude, G. F. V. (2003). Hepatocyte growth factor/scatter factor mediates angiogenesis through positive VEGF and negative thrombospondin 1 regulation. Proceedings of the National Academy of Sciences, 100(22), 12718–12723. 10.1073/pnas.2135113100

Zhou, Yanchen, Penny, H. L., Kroenke, M. A., Bautista, B., Hainline, K., Chea, L. S., … Mytych, D. T. (2022). Immunogenicity assessment of bispecific antibody-based immunotherapy in oncology. Journal for Immunotherapy of Cancer, 10(4), e004225. 10.1136/jitc-2021-004225

Zhou, Yang, Richmond, A., and Yan, C. (2023). Harnessing the Potential of CD40 Agonism in Cancer Therapy. Cytokine & Growth Factor Reviews. 10.1016/j.cytogfr.2023.11.002

